# Individual contralesional recruitment in the context of structural reserve in early motor reorganization after stroke

**DOI:** 10.1101/2024.04.06.24304832

**Authors:** Maike Mustin, Lukas Hensel, Gereon R. Fink, Christian Grefkes, Caroline Tscherpel

## Abstract

The concept of structural reserve in stroke reorganization assumes that the relevance of the contralesional hemisphere strongly depends on the brain tissue spared by the lesion in the affected hemisphere. Recent studies, however, have indicated that the contralesional hemisphere’s impact exhibits region-specific variability with concurrently existing maladaptive and supportive influences. This challenges traditional views, necessitating a nuanced investigation of contralesional motor areas and their interaction with ipsilesional networks.

Our study focused on the functional role of contralesional key motor areas and lesion-induced connectome disruption early after stroke.

Online TMS data of twenty-five stroke patients was analyzed to disentangle interindividual differences in the functional roles of contralesional primary motor cortex (M1), dorsal premotor cortex (dPMC), and anterior interparietal sulcus (aIPS) for motor function. Connectome-based lesion symptom mapping and corticospinal tract lesion quantification were used to investigate how TMS effects depend on ipsilesional structural network properties.

At group and individual levels, TMS interference with contralesional M1 and aIPS but not dPMC led to improved performance early after stroke. At the connectome level, a more disturbing role of contralesional M1 was related to a more severe disruption of the structural integrity of ipsilesional M1 in the affected motor network. In contrast, a detrimental influence of contralesional aIPS was linked to less disruption of the ipsilesional M1 connectivity.

Our findings indicate that contralesional areas distinctively interfere with motor performance early after stroke depending on ipsilesional structural integrity, extending the concept of structural reserve to regional specificity in recovery of function.

## Introduction

Stroke-induced alterations emerge from the local pathology, i.e. the ischemic lesion, over the lesioned hemisphere to contralesional areas, ultimately affecting network configurations and the functional processing in the entire brain (Krakauer et al., 2012; Ward, 2017). A phenomenon frequently observed in functional MRI studies in stroke is increased neural activity within contralesional motor areas during simple motor tasks (Ferris et al., 2018; Gerloff et al., 2005; Rehme et al., 2011). Furthermore, increased contralesional BOLD activity has been reported to be associated with better post-stroke motor function, suggesting its compensatory effect in the light of the vicariation hypothesis (Hallett et al., 2020; Rehme et al., 2012; Ward et al., 2003). However, there is also evidence for maladaptive contralesional effects, framing the concept of interhemispheric competition (Bütefisch et al., 2008; Grefkes et al., 2008). Accordingly, a decrease of inhibitory influences from the ipsilesional hemisphere is thought to result in a subsequent increase of contralesional neural activation and, in turn, enhanced inhibitory influences exerted by the contralesional hemisphere (Carrera and Tononi, 2014; Duque et al., 2005; Murase et al., 2004).

To reconcile contradicting findings concerning the role of contralesional areas, the concept of structural reserve considers contralesional network alterations depending on ipsilesional network characteristics (Murphy and Corbett, 2009; Pino et al., 2014). Correspondingly, whether the influence of the contralesional hemisphere is compensatory or maladaptive is supposed to depend on the amount of ipsilesional unaffected structures spared by the stroke lesion and, thus, remaining functioning of ipsilesional motor areas and corticospinal tract (CST) (Bertolucci et al., 2018; Cramer, 2008; Pino et al., 2014). In patients with a high structural reserve, competitive inhibition from the contralesional hemisphere is more prominent. In comparison, patients with reduced remaining resources in the ipsilesional hemisphere may rely more on compensatory recruitment of contralesional areas (Brancaccio et al., 2022; Pino et al., 2014).

Online transcranial magnetic stimulation (TMS)-interference probes the contribution of specific cortical regions to motor function post-stroke (Hallett, 2000; Kobayashi and Pascual-Leone, 2003). TMS studies have contributed to our understanding of the role of contralesional brain regions in stroke reorganization, indicating that the contralesional hemisphere’s involvement exhibits regional and temporal variability. While early after stroke, the contralesional primary motor cortex (M1) exerts rather maladaptive influences (Bradnam et al., 2012; Volz et al., 2017), this effect may shift towards a more supportive interaction in later, i.e., chronic stages(Lotze et al., 2006; Takeuchi et al., 2008). For contralesional dorsal premotor cortex (dPMC), a relevant influence was identified only in the chronic stage - albeit uniformly supportive (Johansen-Berg et al., 2002; Tscherpel et al., 2020). Furthermore, the impact of posterior parietal areas, i.e., contralesional intraparietal sulcus (aIPS), seems less vulnerable to these temporal dynamics with a rather generally detrimental role for post-stroke motor function (Hensel et al., 2020; Tscherpel et al., 2020).

The interregional differences outlined above challenge the concept of structural reserve, as they instead indicate that contralesional recruitment depends on region-specific interactions with distinct ipsilesional network configurations. In other words, for impacting on the functional relevance of the contralesional hemisphere, the structural reserve may more likely be subject to the regional specificity of different contralesional areas.

Therefore, we aimed to elaborate on previously identified effects of contralesional M1, dPMC, and aIPS in a pooled sample of two methodically equivalent online TMS studies (Hensel et al., 2020; Tscherpel et al., 2020) in relation to ipsilesional structural network resources. To delineate the structural reserve, i.e., the unaffected structures in the ipsilesional hemisphere and the CST, connectome lesion symptom mapping (CLSM) was employed to derive the lesion-induced disconnection severity of the ipsilesional motor network and the specific disintegration of ipsilesional M1 within the ipsilesional motor network (Bowren et al., 2022; Griffis et al., 2021). Likewise, CST integrity was determined by probabilistic and z-axis corrected tract-specific lesion mapping(Feng et al., 2015; Zhu et al., 2010).

Given the region-specificity of TMS effects observed in previous studies, we hypothesized that distinct properties of the ipsilesional structural network pathology differentially contribute to the functional relevance of contralesional motor areas.

## Material and Methods

### Data Pooling

A pooled dataset combining Tscherpel et al. (2020c) and Hensel et al. (2020) was used, sharing a comparable cohort and similar experimental design with respect to the performed finger-tapping task and online TMS interference. No statistical differences were observed between the pooled samples regarding demographics (age, sex), clinical data (days post-stroke, National Institutes of Health Stroke Scale (NIHSS)) or cortical excitability (RMT) (please see Supplementary Material I). A significant difference was found for the average tapping speed in the sham condition, resulting from a different predefined height reference (see below *Index finger-tapping task*) for the tapping movements (patients: F(2,22)=22.30, p<.001, η²=0.11; controls: F(2,21)=41.23, p<.001, η²=0.13) and which was corrected via sham normalization (patients: F(2,22)=2.30, p=0.143); controls: (F(2,21)=0.13, p=0.821).

### Participants

Twenty-five stroke patients (age=67.7 ± 11.6 standard deviation (SD), range: 52-82 years; 8 females; 2 left-handed) with unilateral mild to moderate motor impairment (NIHSS=2.6 ± 2.0) were recruited from the Department of Neurology of the University Hospital of Cologne (Table 1) within the first days after stroke (days post-stroke=3.96 ± 1.97). The following inclusion criteria were applied in both studies (Hensel et al., 2020; Tscherpel et al., 2020): (i) 40-90 years of age, (ii) first-ever ischemic stroke with a unilateral lesion verified by diffusion-weighted MRI (DWI), (iii) study participation within <10 days post-stroke onset, (iv) unilateral upper limb motor deficit, (v) no contraindication to TMS (vi) no cerebral hemorrhage, (vii) no other neurological deficits, i.e., aphasia, apraxia, neglect, visual field deficits, (viii) no pre-existing neurodegenerative or neuroinflammatory disease, (ix) no significant pre-existing musculoskeletal impairment, (x) MRI performed within <48 hours after stroke onset. Two patients of the original sample of Hensel et al. (2020) had to be excluded since the time point of MRI was not performed within the first 48 hours and thus not comparable with the rest of the cohort for the present research question.

23 healthy subjects which were matched to the patient cohort with respect to age, sex and handedness resulted from the two datasets (age=64.2 ± 8.1, range: 50-89 years; p=0.23; 8 females, p=0.92; 1 left-handed). The One healthy subject had to be excluded due to extreme values exceeding two standard deviations of TMS effects, resulting in 22 data sets. Both studies were conducted under the Declaration of Helsinki and approved by the local ethics committee at the University Hospital of Cologne. Prior to the inclusion, all participants provided written informed consent.

### Experimental Design

The single-blinded, sham-controlled experiment targeted three brain regions: the contralesional primary motor cortex (M1), dorsal premotor cortex (dPMC), and anterior intraparietal sulcus (aIPS) (Hensel et al., 2020; Tscherpel et al., 2020) (Figure 1). A placebo stimulation over vertex served as a control condition (Sham). Neuro-navigated 10Hz repetitive TMS (rTMS) was performed time-locked to a finger-tapping task (online TMS interference) to interfere with the ongoing brain activity and investigate the impact of the abovementioned areas of the extended motor network on task performance (Lefaucheur et al., 2020, 2017, 2014; Rossi et al., 2021, 2009). TMS was administered with 90% of the resting motor threshold (RMT). For a detailed description of the TMS protocol, RMT assessment, and TMS site definition, please see Supplementary Material II. The stimulation intensities used for TMS interference did not differ between patients (51.0% maximum stimulator output (MSO) ± 12.3%) and healthy controls (50.4% MSO ± 13.8%, p=0.87).

**Figure 1.**
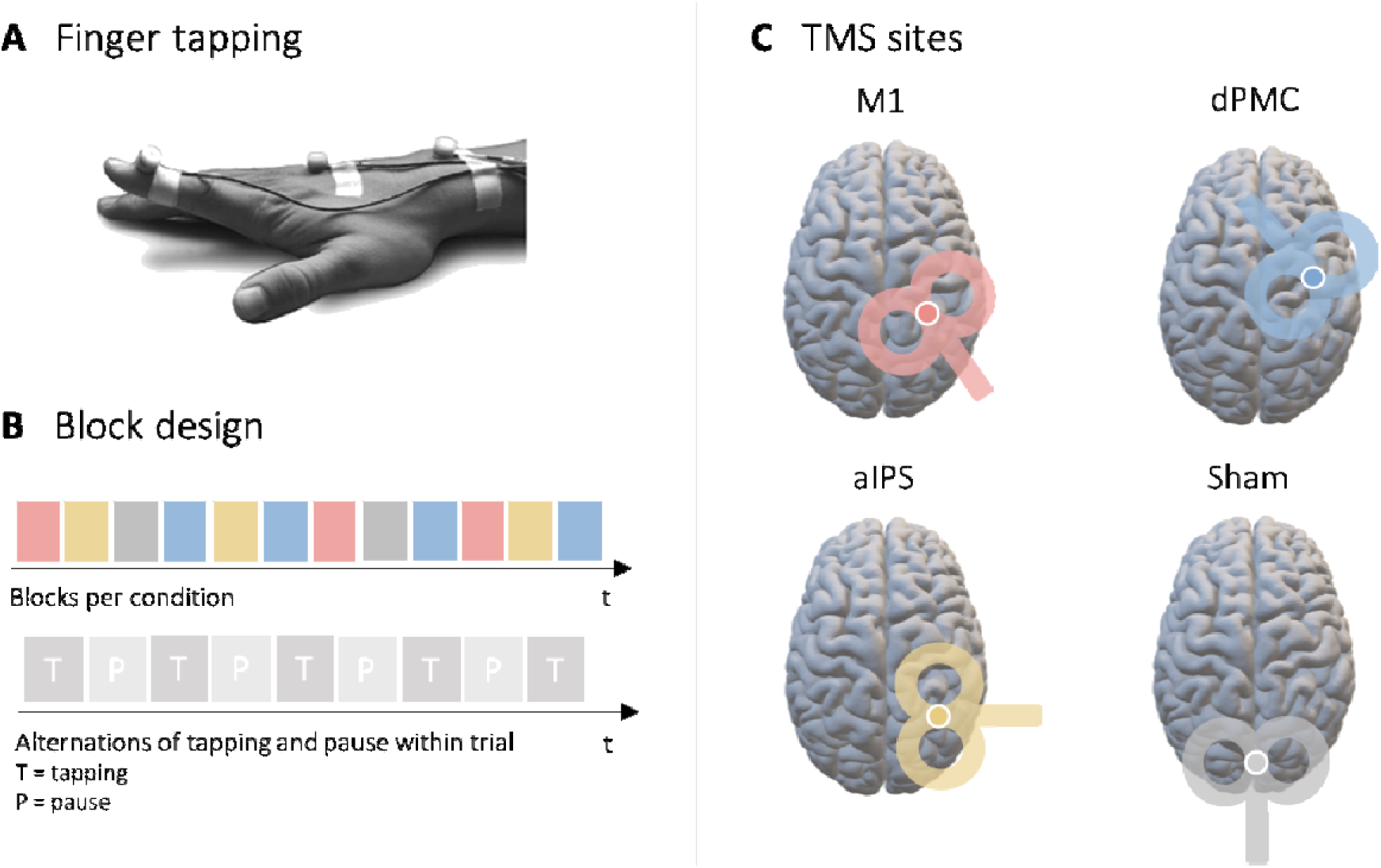
Experimental setup. **A** During applying 10Hz online TMS interference, all subjects performed a simple finger-tapping task in response to a visual cue. **B** The experiment was set up in a pseudo-randomized block design addressing each target region thrice. Each trial consisted of alternating tapping and rest intervals. **C** The three contralesional target regions comprise the primary motor cortex (M1), the dorsal premotor cortex (dPMC), and the anterior intraparietal sulcus (aIPS). The parieto-occipital vertex served as a control condition (Sham).

### Index finger-tapping task and 3D ultrasound movement kinematics

Participants performed repetitive finger-tapping movements with their index finger in response to a visual cue (Figure 1). Both studies applied an alternating block design that rotated through the different target regions, i.e., contralesional M1, dPMC, and IPS. Motor performance was recorded using an ultrasound-based 3D kinematic system (Zebris CMS20 motion analyzer system) tracking the position of the index finger in the mediolateral, anteroposterior, and vertical planes, and subsequently analyzed offline (Hensel et al., 2020; Tscherpel et al., 2020). To assess the stimulation effect, we employed the relative difference (%) of the maximum finger tapping velocity between the stimulation site of interest, i.e., contralesional M1, dPMC, or aIPS, and the Sham stimulation for further analysis. Please see Supplementary Material III for a detailed description of task, recording, and analysis procedures.

### Lesion mapping

For all included patients, standard MR images (1.5T MRI scanner, see Supplementary material IV for further specifications) were taken as part of the clinical diagnostic procedure within 48h after stroke onset. An individual lesion map was manually drawn for each patient based on the DWI image (see Supplementary Material IV for the preprocessing). Standard lesion characteristics were calculated, i.e., the number of affected voxels and lesion volume.

### Voxel lesion symptom mapping

We conducted a voxel lesion symptom mapping (VLSM) using the non-parametric mapping (NPM) software package (Rorden et al., 2007). Only voxels that were damaged in at least 15% of the patients were included in the analysis. We corrected for multiple comparisons using the non-parametric permutation tests (1000 permutations) recommended for medium sample sizes (Medina et al., 2010). Further, we set a significance level of 0.05 FDR corrected as a threshold for voxel consideration.

### Tract-specific lesion mapping

The structural integrity of the CST was quantified using a z-axis corrected probabilistic mapping procedure (Feng et al., 2015; Zhu et al., 2010), accounting for individual variations in brain anatomy and lesion location. Therefore, the calculation of the probabilistic CST lesion overlap (Chenot et al., 2019) was determined separately for each slice with an individual weighting factor according to the height of the lesion along the z-axis, adjusting for the width of the CST on the corresponding slice (Figure 2A). Maximum lesion extent was defined as the greatest extent of the infarct (number of voxels) within the CST relative to the full CST extent in the z-plane. The detailed implementation can be found in the Supplementary Material V.

**Figure 2.**
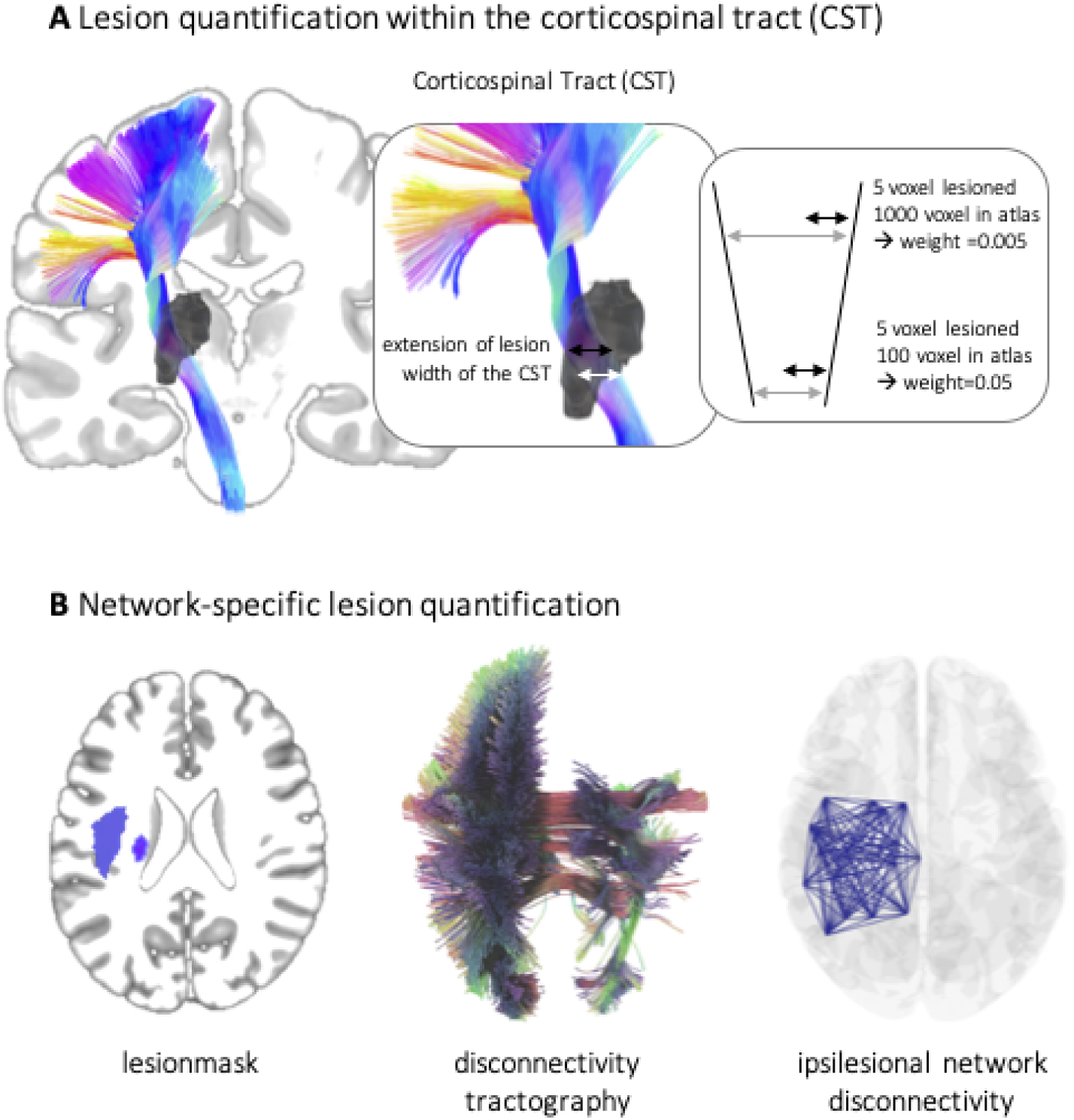
Lesion symptom mapping. A A probabilistic mapping procedure was utilized to assess the structural integrity of the corticospinal tract corrected for the width of the corticospinal tract on the z-axis. This method involved characterizing the extent of the lesion for each height of the tract and weighting it by the individual width at the level of each respective slice. B White matter disconnectivity topographies were created by projecting the individual brain lesion on the HCP-842 human connectome atlas. Based on this overlap between lesion mask and tractography data, a matrix of estimated fiber tract damage was estimated and, thus, derived a lesion-specific disconnectivity profile for each patient.

### Network-specific disconnectivity mapping

A network-specific disconnectivity mapping approach was used to quantify the structural disruption within the ipsilesional motor network (Figure 2B). Using the Lesion Quantification Toolkit (LQT)^32^, white matter disconnectivity matrices were created for each patient by projecting the individual lesion map onto the HCP-842 human connectome atlas (Glasser et al., 2016). By means of a normative database of white matter tractography from healthy subjects, the LQT toolbox identifies and maps the white matter tracts passing through each lesion. By calculating the overlap between the lesion mask and tractography data, the toolbox produces a profile of disrupted white matter pathways specific to each lesion, i.e. a lesion-specific disconnectivity profile. This approach enabled the determination of the disruption of all fiber streamlines connecting cortical parcels (Schaefer et al., 2017), characterizing the lesion-induced damage of gray matter and white matter tracts. Here, a subdivision of an initial parcellation of 100 parcels (Schaefer et al., 2017) focusing on an extended motor network was utilized (see Supplementary Materials V for a list of included parcels)

A graph-theoretical approach was used to summarize the structural loss, focusing on the average node strength in the disconnectivity matrix. This approach measured the amount of disconnection between specific brain regions (parcels) within the ipsilesional motor network. This method allowed to quantify the disconnectivity of (i) the whole network, (ii) the ipsilesional motor network and (iii) particularly the ipsilesional M1’s disconnectivity with the network (details in Supplementary Material V).

### Statistical Analyses

Statistical analyses were performed using the SciPy Stats package (version 1.9.3) in Python (version 3.11). TMS effects were compared between stroke patients and healthy participants for each target region. In case of violation of normal distribution non-parametric test was chosen. To account for the individual heterogeneity of the TMS effect, we assessed the distribution of subjects exhibiting a statistically relevant positive or negative response to TMS. Given that the data of both groups approximated left-skewed distributions centered close to zero and assuming a slight to moderate deviation from normal distribution, a response was considered statistically relevant when exceeding ±1.5 standard errors of the aggregated mean (SEM) of the sham condition. The proportion of responders and non-responders in both groups was compared by Fisher’s exact test.

In order to infer the association between the ipsilesional structural network alterations and the functional relevance of contralesional motor regions, Spearman correlations were performed linking the lesion and disconnectivity measures to region-specific TMS effects in TMS responsive patients. All statistical tests were two-tailed, a significance level of p<0.05 was considered statistically significant, and FDR-correction was used to control for multiple testing.

## Results

### TMS effects early after stroke

Comparing the sham-normalized TMS-induced performance changes, significant group differences in tapping performance were found for contralesional M1 (Z=-2.26, p=0.026, FDR-corrected) and contralesional aIPS (Z=-2.06, p=0.026, FDR-corrected) but not for dPMC (p=0.099) stimulation, indicating a behavioral improvement upon TMS interference with the two former regions in stroke patients early after stroke (Figure 3).

**Figure 3.**
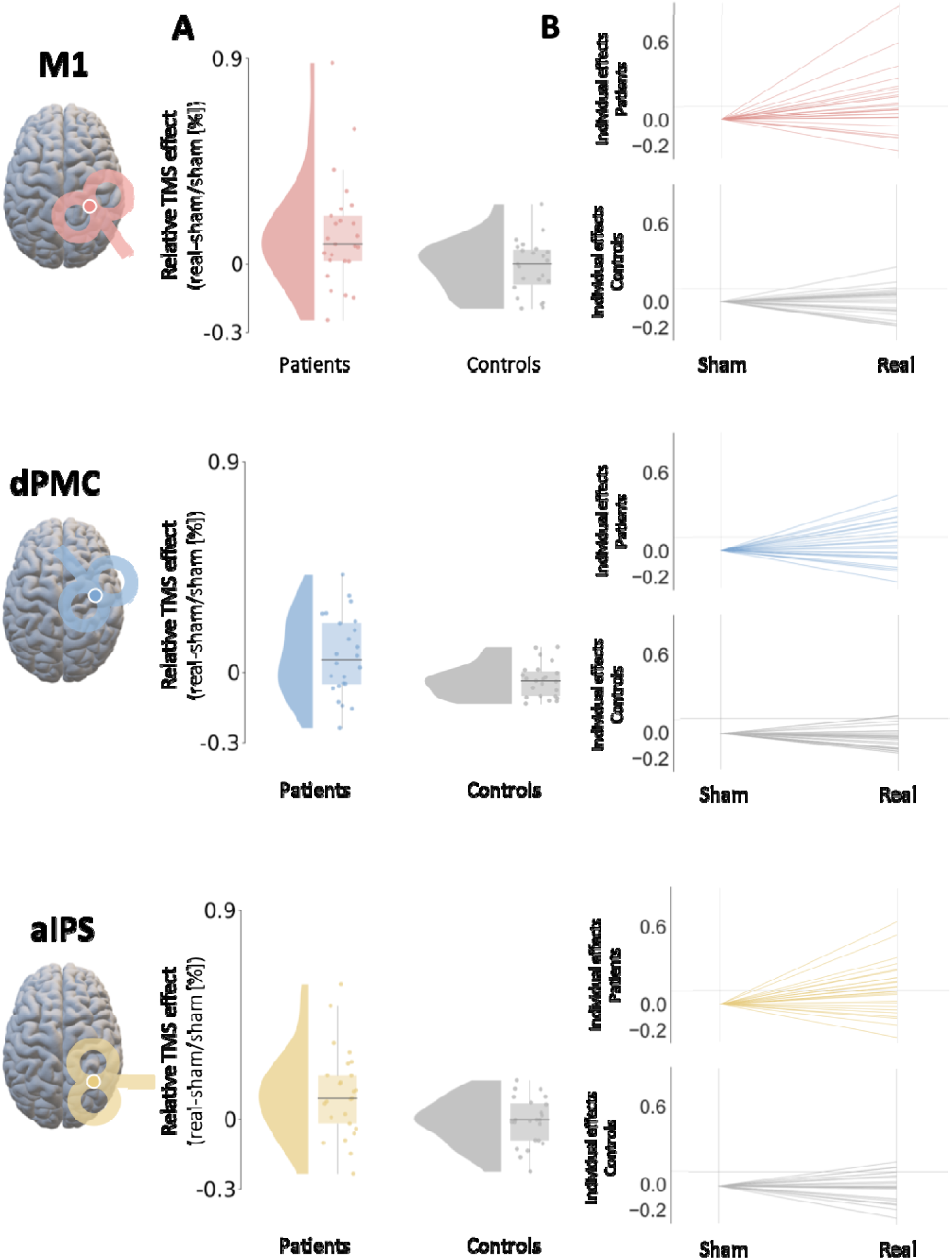
Group comparisons between stroke patients and healthy control subjects and individual data points. **A** A significant difference in finger-tapping velocity between stroke patients and healthy control subjects was found for the stimulation of contralesional M1 and aIPS. Accordingly, patients early after stroke demonstrated an improvement in tapping performance upon contralesional online TMS interference. An equivalent group difference concerning dPMC could not be detected in the present sample. **B** It was demonstrated that the group of stroke patients comprised a significantly higher proportion of positive responders for the M1 and aIPS stimulation but not for dPMC. Patient data are shown in color while healthy controls are shown in gray.

We next analyzed the individual responses, i.e., motor performance improvement or deterioration upon TMS interference at an individual level. Accordingly, for ipsilesional M1, 11 out of 25 patients (44%) but only 2 out of 23 control subjects (8.7%) showed a positive response. For dPMC stimulation, 10 out of 25 patients (40%) demonstrated a positive response, and 3 out of 23 (13%) healthy participants. Finally, when interfering with aIPS activity, 12 out of 25 patients (48%) exhibited a positive effect, while only 3 out of 23 controls (13%) featured similar improvements. Correspondingly, Fisher’s exact test revealed a significantly higher proportion of positive TMS responders in patients for M1 (T=0.12, p=0.048, OR=0.41, FDR-corrected) and aIPS stimulation (T=0.16, p=0.048, OR=0.39, FDR-corrected). In contrast, dPMC stimulation effects (p=0.050) did not pass the statistical threshold (Figure 3). By contrast, there were no comparable effects for individual responses concerning the deterioration of motor performance. Hence, a TMS-induced motor improvement upon contralesional M1 and aIPS interference in patients at the group level was accompanied by a greater proportion of individual positive TMS responses in patients following contralesional M1 and aIPS stimulation.

### General lesion characteristics

The lesion overlap indicated one peak located at the posterior limb of the internal capsule (MNI coordinates: -27, -13, 6) and one in the medioventral part of the pons (MNI coordinates: -4, -20, -30) (Figure 4A). However, lesion volume did not differ between TMS responders and non-responders (p>0.4). Likewise, we did not find differences between the group of TMS responders and the rest of the patients for demographic, behavioral, or clinical data (p>0.3).

**Figure 4.**
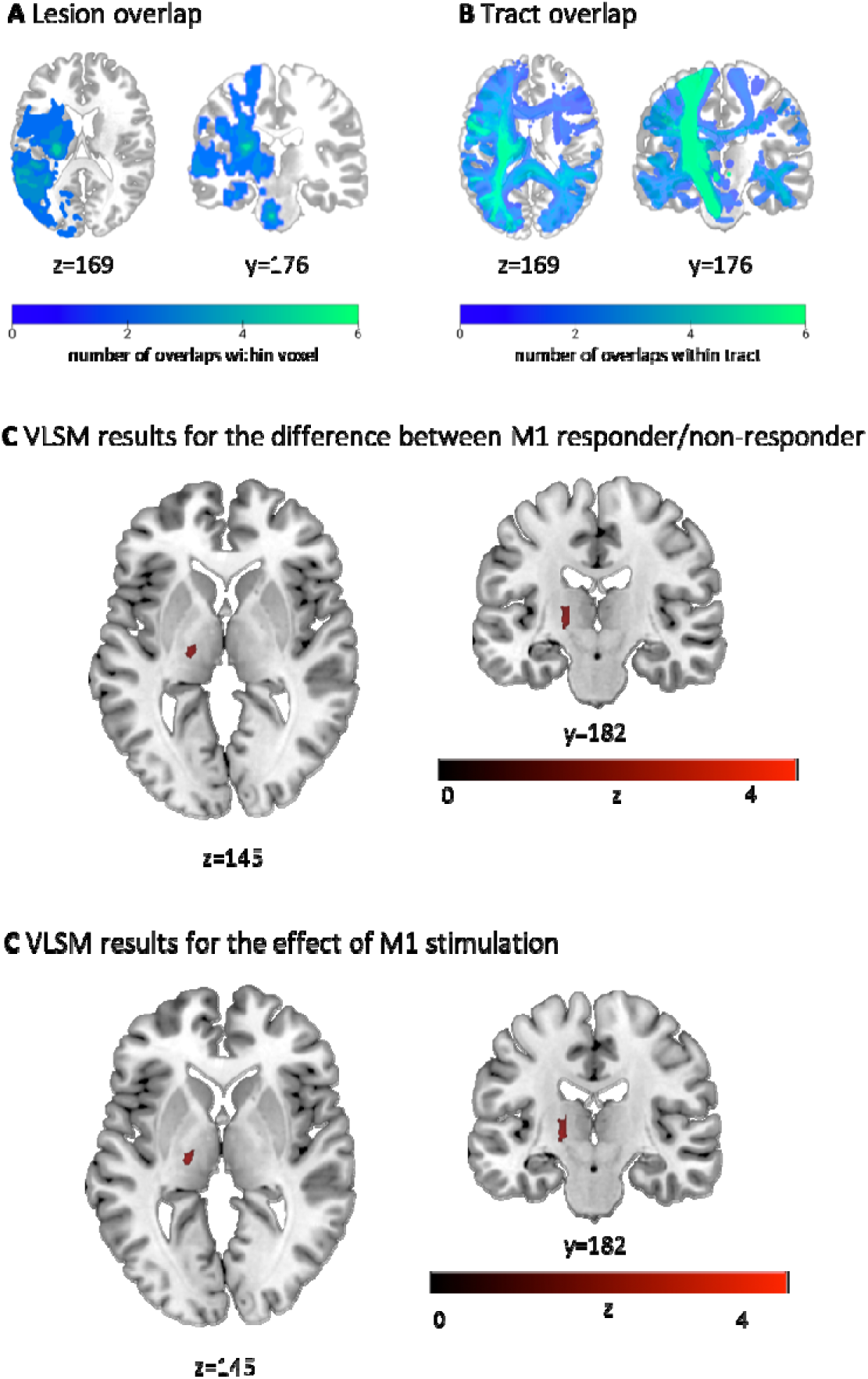
Distributions and effects of lesion location. A The lesion overlap of all patients indicated two peaks: one in the posterior limb of the internal capsule and one in the medioventral part of the pons. B The tract overlap showed a dominant overlap with CST. Please note that lighter colors represent a higher incidence of overlap. C VLSM result for the difference between responders and non-responders upon M1 stimulation. D The VLSM analysis per target region indicated that lesion involving the CST, specifically the posterior limb of the internal capsule influencing the TMS response of contralesional M1. Of note, these relationships were not found for contralesional aIPS or dPMC effects, indicating that those may be rather driven by more complex, e.g., network effects rather than by local factors.

### Impact of CST lesion load on TMS responsivity early after stroke

We further investigated the relationship between CST integrity and TMS responses. Within the group of M1 responders, i.e., the proportion of patients featuring a significant behavioral improvement upon TMS interference with M1 at the individual level, a positive association between the TMS-induced improvement of motor performance upon contralesional M1 stimulation and the maximum CST lesion extent (r=0.76, p=0.014, FDR-corrected; n=11) was observed. Hence, a more substantial disruption of CST fibers was linked to a more detrimental effect of the contralesional M1 on motor performance early after stroke (Figure 5A).

**Figure 5.**
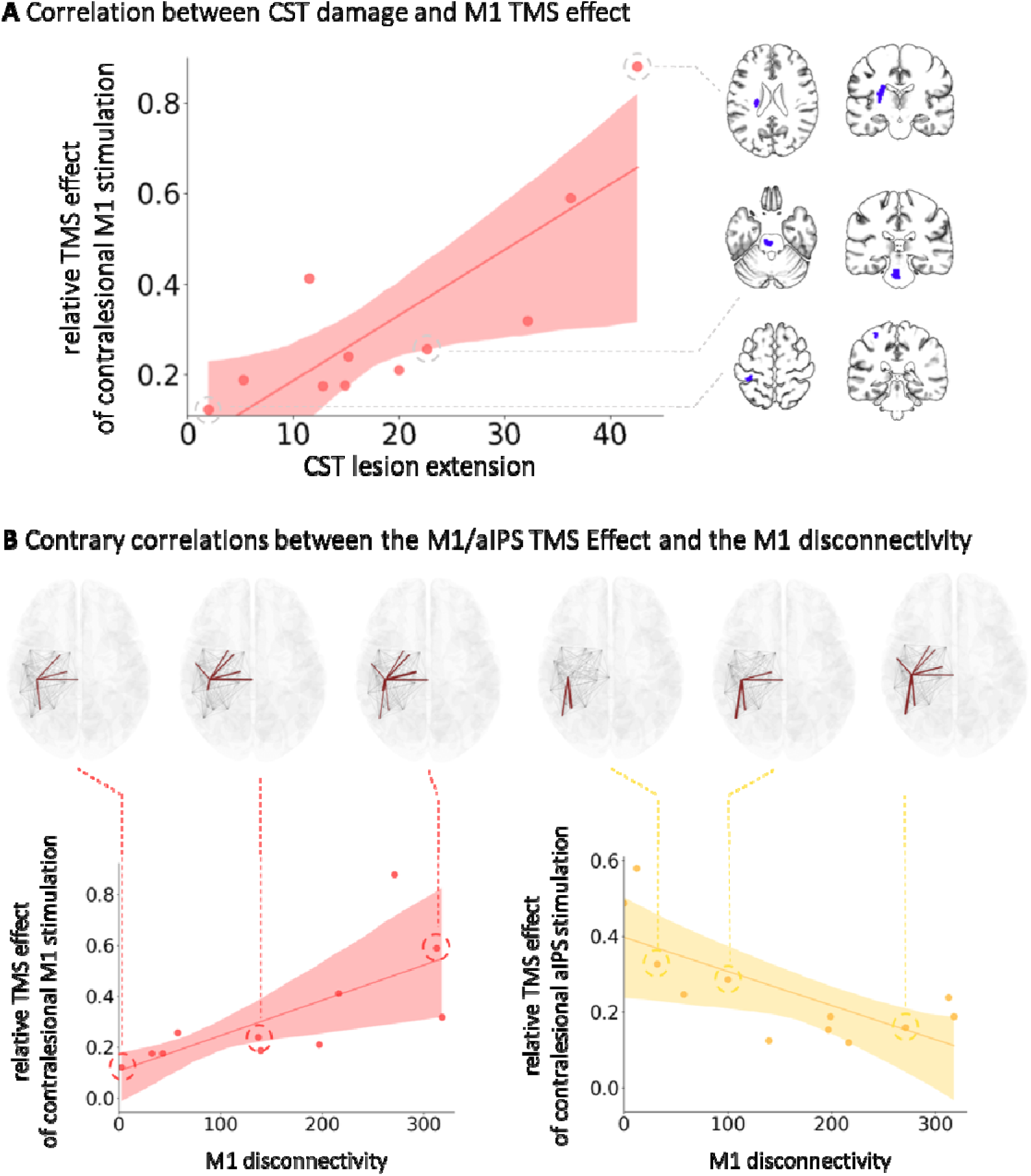
Relationship between ipsilesional network alterations and responsiveness to contralesional TMS. A A positive relationship between the degree of ipsilesional CST lesion load and TMS responsivity to contralesional M1 stimulation was found. B A positive association was found linking the amount of contralesional M1 response to the degree of disconnectivity of the ipsilesional M1. A negative correlation was identified between the contralesional aIPS responses and ipsilesional M1 disconnectivity.

In a linear regression model, a variance explanation of 75% for M1 responsiveness was achieved based on the combined influence of CST lesion extent and the initial upper limb deficit (R^2^=0.75, R^2^ =0.61, p=0.004). In a hierarchical model considering both factors, only the involvement of CST was selected, explaining 67% of the variance (R²=0.67, R²_corr_=0.63, p=0.002), implying that the initial deficit might add to the explanation of the contralesional M1 effect, the contribution of the CST lesion extent, however, was crucial.

In contrast, comparable effects could not be found for aIPS or dPMC stimulation, underpinning the specificity of the interaction of ipsilesional CST affection, motor deficit, and contralesional M1 involvement.

### The relevance of lesion location for TMS responsivity early after stroke

To evaluate the impact of lesion location on TMS responsivity as well as the size of interference effect with contralesional motor areas and, thus, their role for post-stroke motor function, binary and continuous VLSM analyses were conducted for each region of interest. Both yielded significant effects exclusively for M1 stimulation but not aIPS and dPMC (Figure 4C,D). Notably, lesions located in the posterior limb of the internal capsule at the border to the thalamus differentiated between M1 responders and non-responders. Moreover, similar lesion locations were associated with a TMS-induced improvement in motor performance upon contralesional M1, suggesting a detrimental influence of contralesional M1 in case of lesion-induced disruption of the posterior limb of the internal capsule, which further underlines the association between M1 effect and CST integrity reported above. Interestingly, no effect was found for aIPS or dPMC, which suggests that the effects of these two regions may be based on more complex network effects that cannot be explained using local factors.

### Structural reserve of the ipsilesional motor network early after stroke

Based on the hypothesis of structural reserve, we finally tested the relationship between ipsilesional connectome integrity and the effect induced by TMS interference with the two contralesional motor areas of interest. Therefore, TMS responsivity of contralesional M1 and aIPS, i.e., the significant improvement in task performance upon TMS interference with the respective region at the individual patient level, were related to both (i) the averaged node strength and (ii) the M1-specific node strength obtained from the disconnectivity matrix, i.e., the lesion-induced structural damage with respect to white matter tracts of the ipsilesional hemisphere.

Accordingly, no significant associations were found between the more general ipsilesional motor network disconnectivity and the behavioral responsivity upon contralesional M1 or aIPS interference (all p-values>0.4).

By contrast, regarding the disruption of connections specific to ipsilesional M1, opposite dependencies were observed for TMS responsivity of contralesional M1 and aIPS: A positive association was found between the TMS-induced performance improvement upon contralesional M1 interference and the ipsilesional M1 network disruption (r=0.84, p=0.002, FDR-corrected; n=11), indicating that a detrimental influence of contralesional M1 on motor function early after stroke was found explicitly in case of more severe ipsilesional M1 network disruption, i.e., decreased structural reserve (Figure 5B). aIPS responsivity, i.e., improving motor performance upon interference with contralesional aIPS, however, was negatively related to ipsilesional M1 network disruption (r=0.65, p=0.032, FDR-corrected; n=12). Hence, more detrimental influences from contralesional aIPS were particularly found in patients with preserved connections between ipsilesional M1 and the ipsilesional motor network, i.e., greater structural reserve. Thus, albeit TMS interference with both contralesional M1 and contralesional aIPS induced concordant effects on behavioral outcomes, effects on the individual level and their distinct associations with ipsilesional network configurations emphasize the presence of divergent underlying mechanisms at the neural level.

When computing connectivity for ipsilesional aIPS, no significant effects were found neither for TMS effects evoked by contralesional aIPS nor contralesional M1 stimulation (p-values>0.7), underlining the specificity of TMS effects for the ipsilesional M1 connectome. Furthermore, there was no correlation between M1-specific disconnectivity and maximum CST lesion load (r=0.38, p=0.18; n=11), suggesting that these two parameters contain differential information about ipsilesional structural network pathology, such as the functionality of the CST as a descending neural pathway in contrast to cortico-cortical and cortico-subcortical interconnectivity.

## Discussion

We here show that TMS effects on contralesional M1 and contralesional aIPS activity are linked to the degree of network disruption caused by the ischemic lesion early after stroke. To this end, a pooled dataset from two methodologically similar online TMS studies (Hensel et al., 2020; Tscherpel et al., 2020) was utilized to elaborate on the variability of previous findings regarding the recruitment of the contralesional hemisphere early post-stroke. Based on the concept of structural reserve proposing the functional relevance of the contralesional hemisphere in dependence on remaining unaffected structures of the ipsilesional motor system, we used a lesion mapping approach and graph theoretical measures to individually characterize the structural damage, i.e., the lesion-induced disconnectivity. Correspondingly, the relationship between ipsilesional M1 disconnectivity and contralesional TMS effects was opposite for M1 and aIPS, even though at both the group and individual behavioral level, interference with either of the two areas led to an improvement of motor performance of the stroke-affected hand. This observation emphasizes that the individual functional contribution of the contralesional hemisphere to motor function early after stroke can be demonstrated as a function of distinctive region-specific properties of the ipsilesional structural reserve spared by the lesion.

### Motor reorganization in the frontoparietal network early after stroke

A frequently observed phenomenon post-stroke encompasses an increase of neural activity within the contralesional hemisphere during movements of the stroke-affected hand (Grefkes et al., 2008; Rehme et al., 2011). Early post-stroke, this heightened contralesional sensorimotor cortex activity is linked to better recovery (Rehme et al., 2011), evolving into a more lateralized activation in the ipsilesional hemisphere with improved motor function in the chronic phase (Grefkes and Fink, 2014; Rehme et al., 2011). Conversely, persistent bilateral activity in chronic stages is associated with less favorable recovery (Ward et al., 2003).

However, the existing literature portrays conflicting results on the functional implications of increased contralesional activation. While some studies indicate compensatory influences from the contralesional hemisphere, consistent with the hypothesis of vicariation (Rehme et al., 2012; Ward et al., 2003), others describe a detrimental impact of contralesional motor areas in line with the theory of interhemispheric competition (Buetefisch, 2015; Murase et al., 2004). Here, TMS studies have provided insights into the role of local brain activity in stroke recovery (Johansen-Berg et al., 2002; Lotze et al., 2006; Tscherpel et al., 2020; Werhahn et al., 2003), indicating the importance of region- and time-specific aspects in contralesional recruitment in motor recovery and reorganization (Tscherpel et al., 2020; Volz et al., 2017).

Contralesional M1’s involvement varies over time, being maladaptive early post-stroke (Volz et al., 2017) but supportive in chronic stages (Lotze et al., 2006). In line with that, we demonstrate that TMS interference led to an increase in finger-tapping velocity, indicating a detrimental role of contralesional M1 in motor reorganization early after stroke. Notably, in contrast to the results of both original publications, which could not find a significant involvement of contralesional M1 (Hensel et al., 2020; Tscherpel et al., 2020), the pooled analysis expands the results of both by pushing the effect above the threshold with a bigger sample size.

Addressing stroke as a network disorder calls for a more integrated view on interactions within the motor network encompassing premotor and posterior-parietal cortex. For example, several fMRI-studies have demonstrated altered neural activity of contralesional posterior parietal areas and its impact on the ipsilesional M1 in motor reorganization post-stroke (Koch et al., 2007; Rehme et al., 2011; Schulz et al., 2015). Accordingly, additional recruitment of ipsi- and contralesional parietal areas was associated with more significant motor impairment early post-stroke (Rehme et al., 2011). In contrast, better motor function during recovery was linked to an activity decrease in contralesional intraparietal cortex (Ward et al., 2003). TMS studies have previously described a detrimental influence of contralesional aIPS on motor function already in the first weeks after stroke and lasting into the chronic phase (Hensel et al., 2020; Tscherpel et al., 2020). Consistently, the current analysis corroborates these findings.

Several studies have shown that the functional connectivity and activation patterns of contralesional dPMC, critical in the frontoparietal network post-stroke, are associated with motor impairment and recovery after stroke (Bestmann et al., 2010; Johansen-Berg et al., 2002). Specifically, increased activity and connectivity between contralesional dPMC and ipsilesional motor regions have been observed in patients with better motor recovery, suggesting a compensatory mechanism in the chronic phase after stroke (Fridman, 2004; Hensel et al., 2023). These findings are consistent with TMS studies, which have similarly reported a positive influence restricted to the chronic stage of stroke recovery(Tscherpel et al., 2020). In line with that, our analysis did not reveal a significant difference in motor performance between patients and healthy controls following TMS interference early post-stroke

### Integrity of the ipsilesional CST and contralesional TMS responsivity

TMS responses exhibited substantial variability with only around 50% of patients demonstrating a significant improvement under stimulation (Bai et al., 2022; Chen et al., 2014; Hildesheim et al., 2022). However, response variability is not exclusive to TMS but echoes a common pattern of neuromodulating techniques, where responses can vary widely among individuals. For example, the therapeutic outcomes of deep brain stimulation (DBS) present a similar level of heterogeneity (Hitti et al., 2020; Martinez-Ramirez et al., 2018). This has led to an increased focus on connectomic approaches incorporating structural and functional connectivity into treatment planning, enabling personalized therapies (Harita et al., 2022; Hollunder et al., 2022; Menardi et al., 2022; Rajamani et al., 2022). Moreover, also in healthy individuals, TMS-induced neural activity has been shown to be predominantly predicted by the structural integrity of the stimulated network, further highlighting the significance of a network perspective (Momi et al., 2023).

Despite two decades of intensive research on brain reorganization following stroke, data on the functional role of particular brain areas concerning functional or structural network changes in the acute stage after stroke remain scarce. Methodological advances like connectome-based lesion-symptom mapping (CSLM) enable a novel new way of estimating the structural network by projecting lesion maps onto structural standard atlases (Bowren et al., 2022; Griffis et al., 2021).

Moreover, efforts have been undertaken to explain the heterogeneous influences of the contralesional hemisphere on motor reorganization and recovery within the bimodal balance–recovery model and the concept of structural reserve (Pino et al., 2014). According to the theoretical framework of structural reserve, a more profound loss of ipsilesional network structures results in a compensatory recruitment of contralesional areas facilitating motor recovery. By contrast, the contralesional hemisphere exerts a predominantly detrimental impact on a less affected ipsilesional network. In this context, it has to be acknowledged that the current cohort exhibited mild to moderate impairments resulting from rather limited lesions to motor-related fibers in the ipsilesional hemisphere. Therefore, given the relatively preserved ipsilesional structural resources, it could be anticipated that these patients demonstrate a greater vulnerability to disturbing contralesional influences according to the concept of structural reserve.

However, the interregional functional differences within the contralesional hemisphere described above render the notion plausible that structural reserve has to be considered with equivalent regional specificity.

Thus, we further elaborated on the associations between behaviorally overlapping reactivity to TMS interference on contralesional M1 and aIPS and ipsilesional network structures spared by the lesion. Motor deficits after stroke highly depend on the lesion-induced severity of the CST damage (Brouwer and Ashby, 1990; Lotze et al., 2006; Zhu et al., 2010). However, the significance of CST integrity in the context of contralesional involvement has yielded conflicting findings in the literature. While some studies have reported a positive link between ipsilesional CST integrity and contralesional recruitment (DeVetten et al., 2010; Lemon, 2008), others have shown no significant relationship or even a negative association (Bestmann et al., 2010; Stinear et al., 2007). By assessing the functional involvement of the contralesional hemisphere with online TMS interference, our results indicate a positive relationship between ipsilesional CST lesion extent and motor performance improvement upon contralesional M1 interference as an index for a disturbing influence in early motor reorganization. In contrast to the hypothesis of structural reserve, the current results suggest that – at least for the ipsilesional CST and in the present sample of mildly to moderately affected patients with a generally higher structural reserve - the ipsilesional hemisphere is susceptible to maladaptive influences of contralesional M1 in case of less CST integrity. Specifically, this association was observed solely for the contralesional M1 effect and was absent for aIPS. These findings are further underlined by the results of the VLSM analysis. Correspondingly, lesions within the CST, i.e. the posterior limb of the internal capsule at the border to the thalamus, distinguished between M1 responsiveness and non-responsiveness and similar lesion locations were also associated with the extent of M1 effect but explicitly not for aIPS.

### Contralesional significance in the context of ipsilesional network configurations

Previous studies have emphasized the role of interhemispheric structural connections in facilitating the interaction between ipsi- and contralesional structures to enhance motor recovery (Lotze et al., 2006; Ward, 2006). We here underpin the importance of the ipsilesional motor network in the context of structural reserve and contralesional influences. Our findings revealed distinct and opposite relationships between the reserve of the ipsilesional network and the effect of contralesional M1 and aIPS for post-stroke motor performance. Specifically, we found a correlation between ipsilesional M1 disconnectivity and a more disturbing influence of contralesional M1. Likewise, more extensive lesions to the ipsilesional CST were associated with a more disruptive influence of contralesional M1. These converging findings strongly challenge the concept of structural reserve in terms of supportive contralesional influences. Instead, contralesional aIPS exerted a supportive influence for motor performance in patients with stronger ipsilesional M1 disconnectivity, supporting the hypothesis of structural reserve. These opposite effects concerning the influences originating from contralesional areas and their link to ipsilesional network properties vote for including regional specificity in the concept of structural reserve.

Concurrently, the findings emphasize the limitations of simplistic models in comprehensively capturing the complexity of the hemispheric interplay during reorganization after stroke. They further highlight the demand for a more nuanced understanding and individualizable models of the relationship between different contralesional motor regions and their unique contributions to motor recovery.

## Conclusion

In conclusion, the present study’s findings extend the concept of structural reserve by adding the factor region to explain the heterogeneous patterns of contralesional recruitment observed after stroke. Our data emphasize the significance of considering region-specific variations in contralesional recruitment within the context of the structural reserve of the ipsilesional motor network. This highlights the nuanced influence of distinct motor regions and underscores the need for a more personalized approach to understand the mechanisms of motor reorganization after stroke.

## Data availability

The data that support the findings of this study are available from the corresponding author, CT, upon reasonable request.

## Data Availability

All data produced in the present study are available upon reasonable request to the authors

## Acknowledgments

GRF and CG were funded by the Deutsche Forschungsgemeinschaft (DFG, German Research Foundation) – Project-ID 431549029 – SFB 1451 (project C05). We thank Jana Freytag, Stella Ritter, and Katharina Lemberg for technical assistance.

## References

1. Bai, Z., Zhang, J., Fong, K.N.K., 2022. Effects of transcranial magnetic stimulation in modulating cortical excitability in patients with stroke: a systematic review and meta-analysis. J. Neuroeng. Rehabilitation 19, 24. 10.1186/s12984-022-00999-4

2. Bertolucci, F., Chisari, C., Fregni, F., 2018. The potential dual role of transcallosal inhibition in post-stroke motor recovery. Restor. Neurol. Neurosci. 36, 83–97. 10.3233/rnn-170778

3. Bestmann, S., Swayne, O., Blankenburg, F., Ruff, C.C., Teo, J., Weiskopf, N., Driver, J., Rothwell, J.C., Ward, N.S., 2010. The role of contralesional dorsal premotor cortex after stroke as studied with concurrent TMS-fMRI. J Neurosci 30, 11926–11937. 10.1523/jneurosci.5642-09.2010

4. Bowren, M., Bruss, J., Manzel, K., Edwards, D., Liu, C., Corbetta, M., Tranel, D., Boes, A.D., 2022. Post-stroke outcomes predicted from multivariate lesion-behaviour and lesion network mapping. Brain 145, 1338–1353. 10.1093/brain/awac010

5. Bradnam, L.V., Stinear, C.M., Barber, P.A., Byblow, W.D., 2012. Contralesional hemisphere control of the proximal paretic upper limb following stroke. Cereb Cortex 22, 2662–2671. 10.1093/cercor/bhr344

6. Brancaccio, A., Tabarelli, D., Belardinelli, P., 2022. A New Framework to Interpret Individual Inter-Hemispheric Compensatory Communication after Stroke. J. Pers. Med. 12, 59. 10.3390/jpm12010059

7. Brouwer, B., Ashby, P., 1990. Corticospinal projections to upper and lower limb spinal motoneurons in man. Electroencephalogr. Clin. Neurophysiol. 76, 509–519. 10.1016/0013-4694(90)90002-2

8. Buetefisch, C.M., 2015. Role of the Contralesional Hemisphere in Post-Stroke Recovery of Upper Extremity Motor Function. Front Neurol 6, 63–10. 10.3389/fneur.2015.00214

9. Bütefisch, C.M., Weβling, M., Netz, J., Seitz, R.J., Hömberg, V., 2008. Relationship Between Interhemispheric Inhibition and Motor Cortex Excitability in Subacute Stroke Patients. Neurorehabilit. Neural Repair 22, 4–21. 10.1177/1545968307301769

10. Carrera, E., Tononi, G., 2014. Diaschisis: past, present, future. Brain 137, 2408–2422. 10.1093/brain/awu101

11. Chen, M.-H., Huang, L.-L., Lee, C.-F., Hsieh, C.-L., Lin, Y.-C., Liu, H., Chen, M.-I., Lu, W.-S., 2014. A controlled pilot trial of two commercial video games for rehabilitation of arm function after stroke. Clin. Rehabilitation 29, 674–682. 10.1177/0269215514554115

12. Chenot, Q., Tzourio-Mazoyer, N., Rheault, F., Descoteaux, M., Crivello, F., Zago, L., Mellet, E., Jobard, G., Joliot, M., Mazoyer, B., Petit, L., 2019. A population-based atlas of the human pyramidal tract in 410 healthy participants. Brain Struct. Funct. 224, 599–612. 10.1007/s00429-018-1798-7

13. Cramer, S.C., 2008. Repairing the human brain after stroke: I. Mechanisms of spontaneous recovery. Ann. Neurol. 63, 272–287. 10.1002/ana.21393

14. DeVetten, G., Coutts, S.B., Hill, M.D., Goyal, M., Eesa, M., O’Brien, B., Demchuk, A.M., Kirton, A., groups, M. and V. study, 2010. Acute Corticospinal Tract Wallerian Degeneration Is Associated With Stroke Outcome. Stroke 41, 751–756. 10.1161/strokeaha.109.573287

15. Duque, J., Hummel, F., Celnik, P., Murase, N., Mazzocchio, R., Cohen, L.G., 2005. Transcallosal inhibition in chronic subcortical stroke. Neuroimage 28, 940–946. 10.1016/j.neuroimage.2005.06.033

16. Feng, W., Wang, J., Chhatbar, P.Y., Doughty, C., Landsittel, D., Lioutas, V., Kautz, S.A., Schlaug, G., 2015. Corticospinal tract lesion load: An imaging biomarker for stroke motor outcomes. Ann Neurol 78, 860–870. 10.1002/ana.24510

17. Ferris, J.K., Neva, J.L., Francisco, B.A., Boyd, L.A., 2018. Bilateral Motor Cortex Plasticity in Individuals With Chronic Stroke, Induced by Paired Associative Stimulation. Neurorehabilit. Neural Repair 32, 671–681. 10.1177/1545968318785043

18. Fridman, E.A., 2004. Reorganization of the human ipsilesional premotor cortex after stroke. Brain 127, 747–758. 10.1093/brain/awh082

19. Gerloff, C., Bushara, K., Sailer, A., Wassermann, E.M., Chen, R., Matsuoka, T., Waldvogel, D., Wittenberg, G.F., Ishii, K., Cohen, L.G., Hallett, M., 2005. Multimodal imaging of brain reorganization in motor areas of the contralesional hemisphere of well recovered patients after capsular stroke. Electroencephalogr Clin Neurophysiology Electromyogr Mot Control 129, 791–808. 10.1016/0924-980x(95)93143-h

20. Glasser, M.F., Coalson, T.S., Robinson, E.C., Hacker, C.D., Harwell, J., Yacoub, E., Ugurbil, K., Andersson, J., Beckmann, C.F., Jenkinson, M., Smith, S.M., Essen, D.C.V., 2016. A multi-modal parcellation of human cerebral cortex. Nature 536, 171–178. 10.1038/nature18933

21. Grefkes, C., Fink, G.R., 2014. Connectivity-based approaches in stroke and recovery of function. Lancet Neurology 13, 206–216. 10.1016/s1474-4422(13)70264-3

22. Grefkes, C., Nowak, D.A., Eickhoff, S.B., Dafotakis, M., Küst, J., Karbe, H., Fink, G.R., 2008. Cortical connectivity after subcortical stroke assessed with functional magnetic resonance imaging. Ann Neurol 63, 236–246. 10.1002/ana.21228

23. Griffis, J.C., Metcalf, N.V., Corbetta, M., Shulman, G.L., 2021. Lesion Quantification Toolkit: A MATLAB software tool for estimating grey matter damage and white matter disconnections in patients with focal brain lesions. Neuroimage Clin 30, 102639. 10.1016/j.nicl.2021.102639

24. Hallett, M., 2000. Transcranial magnetic stimulation and the human brain. Nature 406, 147–150. 10.1038/35018000

25. Hallett, M., Haan, W. de, Deco, G., Dengler, R., Iorio, R.D., Gallea, C., Gerloff, C., Grefkes, C., Helmich, R.C., Kringelbach, M.L., Miraglia, F., Rektor, I., Strýček, O., Vecchio, F., Volz, L.J., Wu, T., Rossini, P.M., 2020. Human brain connectivity: Clinical applications for clinical neurophysiology. Clin. Neurophysiol. 131, 1621–1651. 10.1016/j.clinph.2020.03.031

26. Harita, S., Momi, D., Mazza, F., Griffiths, J.D., 2022. Mapping Inter-individual Functional Connectivity Variability in TMS Targets for Major Depressive Disorder. Front. Psychiatry 13, 902089. 10.3389/fpsyt.2022.902089

27. Hensel, L., Lange, F., Tscherpel, C., Viswanathan, S., Freytag, J., Volz, L.J., Eickhoff, S.B., Fink, G.R., Grefkes, C., 2023. Recovered grasping performance after stroke depends on interhemispheric frontoparietal connectivity. Brain 146, 1006–1020. 10.1093/brain/awac157

28. Hensel, L., Tscherpel, C., Freytag, J., Ritter, S., Rehme, A.K., Volz, L.J., Eickhoff, S.B., Fink, G.R., Grefkes, C., 2020. Connectivity-Related Roles of Contralesional Brain Regions for Motor Performance Early after Stroke. Cereb Cortex 31, 993–1007. 10.1093/cercor/bhaa270

29. Hildesheim, F.E., Silver, A.N., Dominguez-Vargas, A.-U., Andrushko, J.W., Edwards, J.D., Dancause, N., Thiel, A., 2022. Predicting Individual Treatment Response to rTMS for Motor Recovery After Stroke: A Review and the CanStim Perspective. Front. Rehabilitation Sci. 3, 795335. 10.3389/fresc.2022.795335

30. Hitti, F.L., Ramayya, A.G., McShane, B.J., Yang, A.I., Vaughan, K.A., Baltuch, G.H., 2020. Long-term outcomes following deep brain stimulation for Parkinson’s disease. J. Neurosurg. 132, 205–210. 10.3171/2018.8.jns182081

31. Hollunder, B., Rajamani, N., Siddiqi, S.H., Finke, C., Kühn, A.A., Mayberg, H.S., Fox, M.D., Neudorfer, C., Horn, A., 2022. Toward personalized medicine in connectomic deep brain stimulation. Prog. Neurobiol. 210, 102211. 10.1016/j.pneurobio.2021.102211

32. Johansen-Berg, H., Rushworth, M.F.S., Bogdanovic, M.D., Kischka, U., Wimalaratna, S., Matthews, P.M., 2002. The role of ipsilateral premotor cortex in hand movement after stroke. Proc National Acad Sci 99, 14518–14523. 10.1073/pnas.222536799

33. Kobayashi, M., Pascual-Leone, A., 2003. Transcranial magnetic stimulation in neurology. Lancet Neurol. 2, 145–156. 10.1016/s1474-4422(03)00321-1

34. Koch, G., Olmo, M.F.D., Cheeran, B., Ruge, D., Schippling, S., Caltagirone, C., Rothwell, J.C., 2007. Focal stimulation of the posterior parietal cortex increases the excitability of the ipsilateral motor cortex. J Neurosci 27, 6815–6822. 10.1523/jneurosci.0598-07.2007

35. Krakauer, J.W., Carmichael, S.T., Corbett, D., Wittenberg, G.F., 2012. Getting Neurorehabilitation Right. Neurorehab Neural Re 26, 923–931. 10.1177/1545968312440745

36. Lefaucheur, J.-P., Aleman, A., Baeken, C., Benninger, D.H., Brunelin, J., Lazzaro, V.D., Filipovic, S.R., Grefkes, C., Hasan, A., Hummel, F.C., Jä□askel\”ainen, S.K., Langguth, B., Leocani, L., Londero, A., Nardone, R., Nguyen, J.-P., Nyffeler, T., Oliveira-Maia, A.J., Oliviero, A., Padberg, F., Palm, U., Paulus, W., Poulet, E., Quartarone, A., Rachid, F., Rektorová, I., Rossi, S., Sahlsten, H., Schecklmann, M., Szekely, D., Ziemann, U., 2020. Evidence-based guidelines on the therapeutic use of repetitive transcranial magnetic stimulation (rTMS): An update (2014–2018). Clin Neurophysiol 131, 474–528. 10.1016/j.clinph.2019.11.002

37. Lefaucheur, J.-P., André-Obadia, N., Antal, A., Ayache, S.S., Baeken, C., Benninger, D.H., Cantello, R.M., Cincotta, M., Carvalho, M. de, Ridder, D.D., Devanne, H., Lazzaro, V.D., Filipovic, S.R., Hummel, F.C., Jä□askel\”ainen, S.K., Kimiskidis, V.K., Koch, G., Langguth, B., Nyffeler, T., Oliviero, A., Padberg, F., Poulet, E., Rossi, S., Rossini, P.M., Rothwell, J.C., Schönfeldt-Lecuona, C., Siebner, H.R., Slotema, C.W., Stagg, C.J., Valls-Sole, J., Ziemann, U., Paulus, W., Garcia-Larrea, L., 2014. Evidence-based guidelines on the therapeutic use of repetitive transcranial magnetic stimulation (rTMS). Clin Neurophysiol 125, 2150–2206. 10.1016/j.clinph.2014.05.021

38. Lefaucheur, J.-P., Antal, A., Ayache, S.S., Benninger, D.H., Brunelin, J., Cogiamanian, F., Cotelli, M., Ridder, D.D., Ferrucci, R., Langguth, B., Marangolo, P., Mylius, V., Nitsche, M.A., Padberg, F., Palm, U., Poulet, E., Priori, A., Rossi, S., Schecklmann, M., Vanneste, S., Ziemann, U., Garcia-Larrea, L., Paulus, W., 2017. Evidence-based guidelines on the therapeutic use of transcranial direct current stimulation (tDCS). Clin Neurophysiol 128, 56–92. 10.1016/j.clinph.2016.10.087

39. Lemon, R.N., 2008. Descending Pathways in Motor Control. Annu Rev Neurosci 31, 195–218. 10.1146/annurev.neuro.31.060407.125547

40. Lotze, M., Markert, J., Sauseng, P., Hoppe, J., Plewnia, C., Gerloff, C., 2006. The role of multiple contralesional motor areas for complex hand movements after internal capsular lesion. J Neurosci 26, 6096–6102. 10.1523/jneurosci.4564-05.2006

41. Martinez-Ramirez, D., Jimenez-Shahed, J., Leckman, J.F., Porta, M., Servello, D., Meng, F.-G., Kuhn, J., Huys, D., Baldermann, J.C., Foltynie, T., Hariz, M.I., Joyce, E.M., Zrinzo, L., Kefalopoulou, Z., Silburn, P., Coyne, T., Mogilner, A.Y., Pourfar, M.H., Khandhar, S.M., Auyeung, M., Ostrem, J.L., Visser-Vandewalle, V., Welter, M.-L., Mallet, L., Karachi, C., Houeto, J.L., Klassen, B.T., Ackermans, L., Kaido, T., Temel, Y., Gross, R.E., Walker, H.C., Lozano, A.M., Walter, B.L., Mari, Z., Anderson, W.S., Changizi, B.K., Moro, E., Zauber, S.E., Schrock, L.E., Zhang, J.-G., Hu, W., Rizer, K., Monari, E.H., Foote, K.D., Malaty, I.A., Deeb, W., Gunduz, A., Okun, M.S., 2018. Efficacy and Safety of Deep Brain Stimulation in Tourette Syndrome: The International Tourette Syndrome Deep Brain Stimulation Public Database and Registry. JAMA Neurol. 75, 353. 10.1001/jamaneurol.2017.4317

42. Medina, J., Kimberg, D.Y., Chatterjee, A., Coslett, H.B., 2010. Inappropriate usage of the Brunner–Munzel test in recent voxel-based lesion-symptom mapping studies. Neuropsychologia 48, 341–343. 10.1016/j.neuropsychologia.2009.09.016

43. Menardi, A., Momi, D., Vallesi, A., Barabási, A.-L., Towlson, E.K., Santarnecchi, E., 2022. Maximizing brain networks engagement via individualized connectome-wide target search. Brain Stimul. 15, 1418–1431. 10.1016/j.brs.2022.09.011

44. Momi, D., Wang, Z., Griffiths, J.D., 2023. TMS-evoked responses are driven by recurrent large-scale network dynamics. eLife 12, e83232. 10.7554/elife.83232

45. Murase, N., Duque, J., Mazzocchio, R., Cohen, L.G., 2004. Influence of interhemispheric interactions on motor function in chronic stroke. Can J Neurological Sci J Can Des Sci Neurologiques 55, 400–409. 10.1017/s0317167100000196

46. Murphy, T.H., Corbett, D., 2009. Plasticity during stroke recovery: from synapse to behaviour. Nat Rev Neurosci 10, 1–13. 10.1038/nrn2735

47. Pino, G.D., Pellegrino, G., Assenza, G., Capone, F., Ferreri, F., Formica, D., Ranieri, F., Tombini, M., Ziemann, U., Rothwell, J.C., Lazzaro, V.D., 2014. Modulation of brain plasticity in stroke: a novel model for neurorehabilitation. Nat Rev Neurol 10, 1–13. 10.1038/nrneurol.2014.162

48. Rajamani, N., Horn, A., Hollunder, B., 2022. Connectomic Deep Brain Stimulation 527–542. 10.1016/b978-0-12-821861-7.00009-9

49. Rehme, A.K., Eickhoff, S.B., Rottschy, C., Fink, G.R., Grefkes, C., 2012. Activation likelihood estimation meta-analysis of motor-related neural activity after stroke. Neuroimage 59, 2771–2782. 10.1016/j.neuroimage.2011.10.023

50. Rehme, A.K., Fink, G.R., Cramon, D.Y. von, Grefkes, C., 2011. The role of the contralesional motor cortex for motor recovery in the early days after stroke assessed with longitudinal FMRI. Cereb Cortex 21, 756–768. 10.1093/cercor/bhq140

51. Rorden, C., Bonilha, L., Nichols, T.E., 2007. Rank-order versus mean based statistics for neuroimaging. Neuroimage 35, 1531–1537. 10.1016/j.neuroimage.2006.12.043

52. Rossi, S., Antal, A., Bestmann, S., Bikson, M., Brewer, C., Brockmöller, J., Carpenter, L.L., Cincotta, M., Chen, R., Daskalakis, J.D., Lazzaro, V.D., Fox, M.D., George, M.S., Gilbert, D., Kimiskidis, V.K., Koch, G., Ilmoniemi, R.J., Lefaucheur, J.P., Leocani, L., Lisanby, S.H., Miniussi, C., Padberg, F., Pascual-Leone, A., Paulus, W., Peterchev, A.V., Quartarone, A., Rotenberg, A., Rothwell, J., Rossini, P.M., Santarnecchi, E., Shafi, M.M., Siebner, H.R., Ugawa, Y., Wassermann, E.M., Zangen, A., Ziemann, U., Hallett, M., 2020, T. basis of this article began with a C.S. from the I.W. on “Present, Future of TMS: Safety, Ethical Guidelines”, Siena, October 17-20, 2018, updating through April, 2021. Safety and recommendations for TMS use in healthy subjects and patient populations, with updates on training, ethical and regulatory issues: Expert Guidelines. Clin. Neurophysiol. 132, 269–306. 10.1016/j.clinph.2020.10.003

53. Rossi, S., Hallett, M., Rossini, P.M., Pascual-Leone, A., 2009. Safety, ethical considerations, and application guidelines for the use of transcranial magnetic stimulation in clinical practice and research. Clin Neurophysiol 120, 2008–2039. 10.1016/j.clinph.2009.08.016

54. Schaefer, A., Kong, R., Gordon, E.M., Laumann, T.O., Zuo, X.-N., Holmes, A.J., Eickhoff, S.B., Yeo, B.T.T., 2017. Local-Global Parcellation of the Human Cerebral Cortex from Intrinsic Functional Connectivity MRI. Cereb Cortex 28, 3095–3114. 10.1093/cercor/bhx179

55. Schulz, R., Koch, P., Zimerman, M., Wessel, M., Bönstrup, M., Thomalla, G., Cheng, B., Gerloff, C., Hummel, F.C., 2015. Parietofrontal motor pathways and their association with motor function after stroke. Plos One 138, 1949–1960. 10.1371/journal.pone.0052727

56. Stinear, C.M., Barber, P.A., Smale, P.R., Coxon, J.P., Fleming, M.K., Byblow, W.D., 2007. Functional potential in chronic stroke patients depends on corticospinal tract integrity. Brain 130, 170–180. 10.1093/brain/awl333

57. Takeuchi, N., Tada, T., Toshima, M., Chuma, T., Matsuo, Y., Ikoma, K., 2008. Inhibition of the unaffected motor cortex by 1 Hz repetitive transcranical magnetic stimulation enhances motor performance and training effect of the paretic hand in patients with chronic stroke. J. Rehabilitation Med. 40, 298–303. 10.2340/16501977-0181

58. Tscherpel, C., Hensel, L., Lemberg, K., Vollmer, M., Volz, L.J., Fink, G.R., Grefkes, C., 2020. The differential roles of contralesional frontoparietal areas in cortical reorganization after stroke. Brain Stimul. 13, 614–624. 10.1016/j.brs.2020.01.016

59. Volz, L.J., Vollmer, M., Michely, J., Fink, G.R., Rothwell, J.C., Grefkes, C., 2017. Time-dependent functional role of the contralesional motor cortex after stroke. Neuroimage Clin 16, 165–174. 10.1016/j.nicl.2017.07.024

60. Ward, N.S., 2017. Restoring brain function after stroke - bridging the gap between animals and humans. Nat Rev Neurol 13, 244–255. 10.1038/nrneurol.2017.34

61. Ward, N.S., 2006. Motor system activation after subcortical stroke depends on corticospinal system integrity. Brain 129, 809–819. 10.1093/brain/awl002

62. Ward, N.S., Brown, M.M., Thompson, A.J., Frackowiak, R.S.J., 2003. Neural correlates of motor recovery after stroke: a longitudinal fMRI study. Brain 126, 2476–2496. 10.1093/brain/awg245

63. Werhahn, K.J., Conforto, A.B., Kadom, N., Hallett, M., Cohen, L.G., 2003. Contribution of the ipsilateral motor cortex to recovery after chronic stroke. Ann Neurol 54, 464–472. 10.1002/ana.10686

64. Zhu, L.L., Lindenberg, R., Alexander, M.P., Schlaug, G., 2010. Lesion Load of the Corticospinal Tract Predicts Motor Impairment in Chronic Stroke. Stroke 41, 910–915. 10.1161/strokeaha.109.577023

